# The value of freedom: extending the evaluative space of capability

**DOI:** 10.1101/2022.06.29.22277019

**Authors:** Jasper Ubels, Karla Hernandez-Villafuerte, Erica Niebauer, Michael Schlander

**Affiliations:** Division of Health Economics, German Cancer Research Center (DKFZ), Heidelberg, Germany; Mannheim Medical Faculty, University of Heidelberg, Mannheim, Germany; Alfred-Weber Institute, University of Heidelberg, Heidelberg, Germany

**Author notes:** **Corresponding Author:** Jasper Ubels.

## Abstract

**Introduction:** Developing an instrument with an ambiguous construct can be challenging. With the capability approach, this is argued to be case, since the concept of capability by Sen is ambiguous in respect to the burdens that people experience whilst achieving their capabilities. A potential solution is to develop instruments with a more comprehensive concept of capability, such as the concept ‘option-freedom’. The concept option-freedom stresses the importance of achieving capabilities without impediments. However, this concept has not been operationalized for wellbeing assessment. The aim of this study is to develop a theoretical framework of wellbeing with the concept option-freedom.

**Methods:** A best-fit framework synthesis was conducted with seven papers that report qualitative findings which underpin capability instruments. First, the a-priori concept option-freedom was used to deductively code against. New codes, subthemes and themes were developed inductively when data did not match the a-priori concept.

**Results:** Four themes emerged from the synthesis. (1) Option Wellbeing represents a range of options that need to be satisfied in order for individuals to experience wellbeing. (2) Self-Realization represents that there are experiences in an individual’s live that have value beyond realizing options. (3) Perceived Access to Options represents the perceived ability of individuals to realize freedoms. (4) Perceived Control represents the experience of having control.

**Conclusion:** Developing an instrument with the proposed framework has two benefits. First, it acknowledges the importance of assessing impediments in realizing capabilities for the assessment of wellbeing. Second, the themes form a broad informational base by including themes related to subjective wellbeing. The framework could be used as a broad base on which to assess the value of health technologies. Future research should study the feasibility of implementing the framework for the assessment of wellbeing.

## Introduction

Health technology assessment is the practice of assessing the value of new health technologies in order to inform decision making (1, 2). Some jurisdictions use the Quality Adjusted Life Year (QALY) to assess the value of a health technology. The QALY is a measure that combines both information about health-related quality of life and length of life, by adjusting a life year with the quality of that life year. This quality adjustment is calculated by performing weighted adjustments to scores from instrument that measure changes in health related quality of life. The weights that are used for these adjustment reflect the utility of living (3, 4).

However, it is argued that the impact of health technologies is not limited to improving the health of an individual. Therefore, some scholars argue for the use of the capability approach as a framework to assess the value of health technologies (4, 5). They claim that value should be assessed on a broad informational base that goes beyond quality and length of life, and takes into consideration that what is important for the people themselves.

In this context, the capability approach has emerged as an alternative framework for the development of instruments. Two key concept in the capability approach are functioning and capability. Functionings reflect those things that an individual can do or be, for instance, to be nourished or to be a respected member of the community. Capabilities of individuals represent the combination of opportunities that an individual ‘can do’ or ‘can be’ (6).

This conceptualization of capability approach was developed by Sen (7). However, in terms of operationalizing the approach for the assessment of wellbeing, this concept has two limitations. The first limitation is that this conceptualization is relatively narrow, since it defines capabilities as a kind of positive freedom (8) The burdens that people might experience while achieving these capabilities are not well reflected in this definition (9). The second limitation is that this conceptualization of capability is relatively vague, which poses a challenge for operationalizing the capability approach (9). In the context of wellbeing assessment, this results in difficulties in identifying whether certain elements of wellbeing are more appropriately assessed in terms of capabilities or in terms of other constructs, such as functionings (9).

One solution for these limitations is to develop an instrument with an a-priori conceptualization of capability that is clearer and more comprehensive. Robeyns (8) has proposed such a concept. She argues that a capability is best understood as an ‘option freedom’, which is a concept developed by Pettit (10). Robeyns argues that this definition of freedom more accurately describes capability than other conceptualizations.

According to Pettit, an option freedom consists of (A) options and (B) access to those options. Pettit defined (1) *options* as “the alternatives that an individual is in a position to realize” (10). The characteristics of the options themselves can be quantitative (the amount of options) and qualitative (e.g. diversity of the options) in nature. Further characteristics are the objective and subjective significance of the options to individuals as ways to affect the world. In this respect, some options might be more valuable for one individual than another. The (2) *access* to an option is the possibility to realize an option. The access to an option can be blocked or burdened. A more detailed explanation about the access to options can be found in the appendix.

Conceptualizing capability as an option freedom in the context of instrument development has two advantages. First, the concept of option freedom stresses that a capability can be considered as a freedom that can blocked or burdened. As such, by assessing wellbeing based on this definition, more attention is paid to the blocks and burdens that people might experience whilst achieving their capabilities (9). Second, operationalizing a clear concept of capability facilitates the distinction between the parts of a wellbeing framework that represent capability, and parts that represent other elements that might be relevant for wellbeing assessment besides capability.

The concept of option freedom has not been operationalized yet for measurement. This study aims at proposing a theoretical framework that can be used as the first step to identify domains, which can be used to develop an instrument that assesses the wellbeing of individuals based on the concept ‘option of freedom’.

## Methods

In this study, a best-fit framework synthesis is conducted (11). With this method, a theoretical framework is identified *a-priori*, which is used to code the data against. Data which does not fit the a-priori theoretical model are reinterpreted with thematic analysis techniques. The result of such an analysis is a new or further refined conceptual framework.

Our best-fit framework synthesis followed three general steps: (A) the identification of an a-priori framework or theory; (B) the development of a search strategy to identify studies; and (C) the data analysis and synthesis of a new or updated framework. The enhancing transparency in reporting the synthesis of qualitative research statement was used as a checklist (see appendix table 2) to ensure the current study’s transparency (12).

### (1) Identification of an a-priori framework

A definition of capability is used as an a-priori “lens” for analysis. As mentioned, Sen’s definition is ambiguous, for instance, in respect to the burdens that people might experience whilst achieving their capabilities (8, 9). Therefore, capability is defined as an option freedom (10). The advantage option freedom over other definitions of capability is that freedom is understood as something that cannot only be “externally” blocked (through, for example, laws that limit capabilities), but also be “internally” blocked (through, for example, societal conditioning of women to not follow education, which results in women themselves not wanting to follow education). There are also alternative ways to define capability, however, to evaluate the advantages and disadvantages of all of them would entail a philosophical debate that is beyond the scope of this paper. For a further discussion about the various concepts of freedom and how they relate to capability, see Robeyns (8).

### (2) Search strategy

The papers included in this synthesis are selected from an earlier study where we conducted a full systematic literature review (9). The literature review included the articles that explain how instruments were developed to assess capabilities in the context of wellbeing assessment in the field of health (these articles are hereafter called ‘development papers’). The instruments were identified with a comprehensive pearl growing search strategy (13). Relevant papers were searched for in PubMed and Web of Science. The development papers were reviewed to understand how researchers operationalized the measurement of capability. This was done by examining the concepts of capability used, as well as how those concepts were translated into the themes and questions to create an instrument. Further details about the search strategy can be found in Ubels et al. (9).

For the present analysis, development papers were considered eligible for inclusion when they contained “rich” qualitative data. Articles containing rich qualitative data are those articles that do not only mention the themes measured by the instrument, but also explain how these themes were developed. These themes are then supported with quotes from the participants whose insights where use to develop the themes (hereafter called the participants).

The quality of the identified studies was appraised by JU using the Consolidated Criteria for Reporting Qualitative Research (COREQ) checklist (14). These criteria were not used as a standard to exclude studies, given the discussion around the exclusion of qualitative studies in literature review (15). Rather, the checklist was used by the author to ensure that no important criteria had been missed in the reading of the studies which might influence the development of the framework. A further post-hoc sensitivity analysis was conducted to evaluate if excluding studies with missing or unclear information could have influenced the result of synthesis. This was done by comparing the themes identified in the development papers that provided a complete report according to the COREQ checklist, and the development papers that missed reporting some aspects (15, 16).

### (3) Data synthesis

A best-fit framework synthesis was with the aim that the identified themes could be used as constructs for instrument development. First, data was analyzed by extracting the complete result sections of the development papers to Excel. Second, two a-priori themes, ‘Options’ and ‘Access to Options’, were used to deductively analyze the data from the development papers sequentially. These a-priori themes are based on the concept Option Freedom (10). Data that did not fit the a-priori themes, were inductively analyzed using thematic synthesis methods (11). Out of the inductively analyzed data, new codes and new themes were identified. After defining new codes and themes, the data was again analysed and coded against the newly developed codes and themes. This process was iterative, with the aims to further define the themes and to explore the relationships between the themes. Coding was conducted line by line. Information about the authors who developed the framework and their tasks during the coding process can be found in the appendix. All the illustrating quotations from the in this synthesis are from participants of the studies that are included in this analysis.

## Results

Seven out of the eleven development papers identified by Ubels et al. (9) were eligible for inclusion in the best-fit framework synthesis (17-23). These seven papers form the basis of the content of: (A) a capability instrument that is developed to assess wellbeing in individuals affected by chronic pain, developed by Kinghorn et al. (17), (B) a non-invasive prenatal testing related capability wellbeing questionnaire, developed by Kibel and Vanstone (18), (C) the ICECAP-A (ICEpop CAPability measure for Adults) developed by Al-Janabi et al. (19), (D) the ICECAP-O (ICEpop CAPability measure for Older people) developed by Grewal et al. (20), (E) the ICECAP-SCM (ICECAP Supportive Care Measure) developed by Sutton et al. (21), (F) the women’s capability index developed by Greco et al. (22) and (G) a Diabetes specific instrument for measuring patient reported outcomes and experiences in the Swedish national diabetes register developed by Engström et al. (23). The other four papers included in that review, which concern the development of four other instruments (24-27), did not contain the rich data necessary for a best-fit framework synthesis at the time of writing this article and were therefore excluded from the current study.

Based on the COREQ checklist, some observations can be made. Generally, the studies contained detailed information about the data analysis and the findings. However, the relationship between the interviewers and the participants was unclear in two papers (20, 22). Furthermore, in some papers information about the personal characteristics of the researchers were missing (17, 19, 20, 22). It was thus difficult to assess how the individual backgrounds of the researchers could have potentially influenced the interpretation of the data. A detailed review of the qualitative papers included in the present study can be found in Ubels et al. (9).

### Development of the Model

Four main themes emerged as a result of the synthesis: (1) Option Wellbeing, (2) Self-Realization, (3) Perceived Access to Options, and 4) Perceived Control. (1) Option Wellbeing represents the idea that an individual needs to have access to a range of different options that result in happiness or satisfaction. (2) Self-Realization represents the idea that there are experiences in an individual’s life that have value besides happiness and life satisfaction derived from option realization. These experiences give individuals the sense that their lives are worth living. (3) Perceived Access to Options represents the perceived ability of individuals to access those options that they value. This also includes the perceptions of burdens that people might experience in accessing those options. (4) Perceived Control represents the experience of having control, which is influenced by the balancing act between accessible options and the options an individual values, which might or might not be accessible. The perception of having control is a result of the individual’s capacity to successfully managing burdens in the access to options; managing burdens in a way that the individual can achieve those options that he or she values.

The next sub-sections will provide a description of each themes and their related subthemes. Appendix table 1 shows an overview of the themes and subthemes, with definitions and associated quotes from the development papers. Figure 1 presents the four main themes and their associated subthemes that are identified in this study.

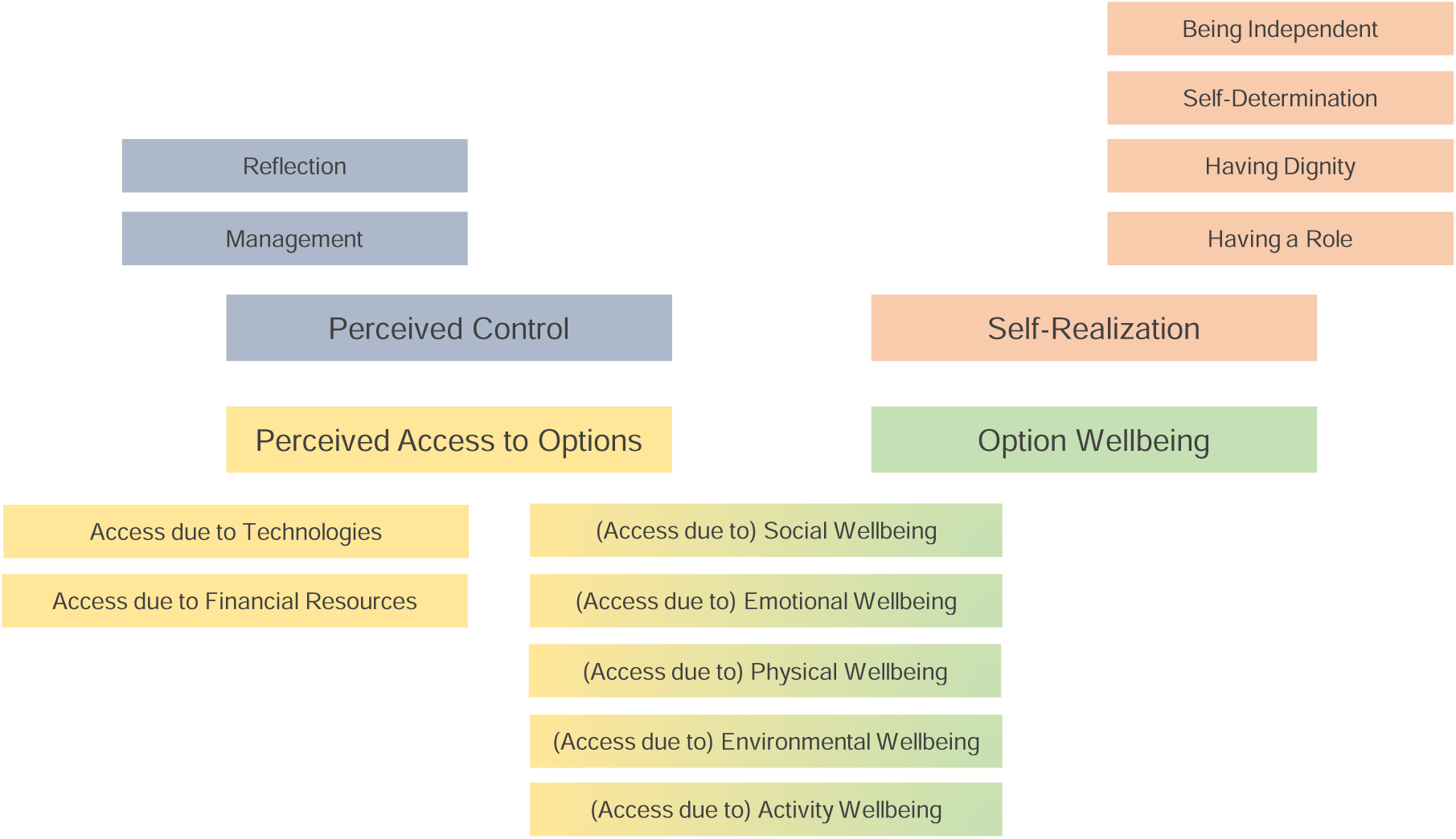

#### 1) Option Wellbeing

The theme Option Wellbeing represents the importance for people have options that are beneficial in terms of happiness or satisfaction. This theme is essentially based on the a-priori concept Options, and reflects that in order for the individual to experience happiness or life satisfaction, certain options need to be realized to what she/he perceives to be an adequate level. If these options cannot be realized to an adequate level, the happiness or satisfaction of individuals is negatively affected. These options could be subdivided in five abstract subthemes: (1.1) Physical Wellbeing, (1.2) Emotional Wellbeing, (1.3) Social Wellbeing, (1.4) Environmental Wellbeing, and (1.5) Activity Wellbeing. A definition of each of these subthemes can be found in appendix table 1.

Depending on the personal characteristics of the participants of the studies, different subthemes appear to be more important in the development papers. For instance, the relevance of achieving a perceived adequate level of Physical Wellbeing had a predominant role when the themes were developed with participants affected by disease (17, 21, 23), with elderly (20), as well as with participants groups for which specific medical technologies are developed (18).

The following quote highlights the negative impact of a lack of Physical Wellbeing:

> *I’m in pain 24-7, whether I’m laying down, in the bath or hanging from the lightshade, I’m in bloody pain (Male, not employed, B), Kinghorn, Robinson (17)*

The importance of Environmental Wellbeing and Activity Wellbeing particularly emerged from four development papers (17, 19, 20, 22). The remaining subthemes, Emotional Wellbeing and Social Wellbeing, were mentioned to be important by the researchers in all the development papers (17-23).

#### 2) Self-Realization

The theme Self-Realization rests on the idea that there are aspects in an individual’s live which have value beyond options that generate happiness or satisfaction. These aspects could be divided in subthemes (2.1) Having a Role, (2.2) Having Dignity, (2.3) Being Independent, and (2.4) Self-Determination. Pursuing these subthemes might even come at a cost of happiness or satisfaction. The importance of being able to experience these aspects was noted in several development papers (17-22).

Having a Role involves more than simply doing certain things for pleasure or satisfaction, as would be the case in the subtheme Activity Wellbeing. Having a Role refers to being able to do those things that provide a sense of worth and identity (17, 19, 20, 22). For example, Kinghorn, Robinson (17) noted that some men perceived themselves to be less masculine, since they could not carry heavy things to help their partners.

> *…you feel inadequate. Well I do, when my missus starts…unloading the car, and I walk into the house and sit down. (M employed, A), Kinghorn, Robinson (17)*

Having Dignity represents the importance of the perceived social standing of an individual in their community. The subtheme is related to the ability of individuals to conduct themselves as beings of worth and be respected by other members of their community. In this context, several papers mentioned the importance of recognition by other people (17-19, 21-23), as well as the ability of people to take care of matters which are considered to be private (21, 22).

> *I’ve got my self-respect, she [carer] doesn’t stand there if I’m having a shower and all that, she just makes sure the windows are covered … we all want our self-respect no matter who we are. (Female, 68 years, PC), Sutton and Coast (21)*

Being Independent is related to participants being free to make their own choices, without being influenced by limitations and having to rely on others to access options. Being Independent can also be understood as the more fundamental perception of individuals to have agency over their own lives, especially in troubled times. This subtheme was particularly highlighted in five papers (17, 19-22).

The subtheme Self-Determination refers to the ability to make valuable choices related to options that are meaningful for the individual. Terminology used by the authors included: (1) the freedom to express oneself without being oppressed (22), (2) the ability of doing certain actions without asking for consent (22), (3) the ability of individuals to achieve goals or move forward in their lives (19) and (4) being able to make choices about aspects that influence their lives (18, 21).

> *“I’m staying here until I get carried away. I’ve worked hard and paid for it, and this is my abode and I’m quite happy with it*.*” (Male, 72 years, GP) Sutton and Coast (21)*

The need to choose, however, was also considered to be overwhelming in some circumstances (18, 21). For instance, when the difficulty of processing the information required to understand and/or weight alternatives overloaded the capacity of individuals to deal with this information, choices were reported to be extremely difficult.

> *[Concerning non-invasive prenatal testing (NIPT)] “Well I think a lot of women don’t really want to think about it but I think they need to understand what the possible outcomes are. Like I think everybody knows that Down’s syndrome is trisomy 21 but there seems to be a lot of vagueness and even a lot of confusion about the testing itself and about what they’re actually looking for*.*” Kibel and Vanstone (18)*

#### 3) Perceived Access to Options

As mentioned, the best-fit framework synthesis started with two a-priori concepts, one of them being Access to options. During the synthesis, the a-priori concept ‘Access to options’ was re-conceptualized into the theme Perceived Access to Options. This change stresses the subjective experienced nature of access to options that was captured by the development papers. The theme Perceived Access to Options represents the perceived ability to access options that are of value to the individuals in regard to their wellbeing. It reflects individuals’ perceptions regarding barriers that exist in their lives.

The same subthemes included in Option Wellbeing can be considered here: Physical Wellbeing (17-23), Emotional Wellbeing (17, 22, 23), Social Wellbeing (17-23), Environmental Wellbeing (17-22) and Activity Wellbeing (17, 19, 20, 22, 23). Additionally, the Perceived Access to Options is also influenced by two other factors, the financial situation of participants and the use of (medical) technologies (17-20, 22, 23). A short definition per subtheme in the context of access to options can be found in appendix table 1.

The subthemes here are mutually interdependent, meaning that burdens or blocks in the access to one option could influence the access to other options. For example, a low level of Physical Wellbeing (e.g. problems with walking associated to chronic pain) might limit the access to options of the subtheme Social Wellbeing, as can be observed in the following quote.

> *‘s sad not daring to go [on a trip]. (*…*) Since it [hypo-glycaemia] is a threat, it feels like a lower quality of life. (*…*) You get a little scared of exposing yourself to situations other than what you are used to. (#17; Woman, 60 years old, Type 2 DM) Engström, Leksell (23)*

When the achievement of one option lead to the limitation or burden the access to other options, a balance between the achievements of options need to be done. For example, an individual’s Physical Wellbeing might improve due to the use of a drug to relieve pain. However, the drug itself could also have a negative impact on Physical Wellbeing through its side effects. This creates a balancing act.

> *“The side-effects can be as bad as the pain itself*.*” (Male, Employed), Kinghorn, Robinson (17)*

#### 4) Perceived Control

The theme Perceived Control represents the importance of people to have a perceived “grasp” over their lives. Authors noted in their development papers the importance of this perception of control for individuals (17-20, 22). This perception of control is defined by its subthemes Management and Evaluation.

The subtheme Management represents the perceived ability of individuals to use particular options in order to reduce the effects of factors that limits the access to other options. In a way it is related to the Perceived Access to Options theme, where the level of achievement in certain options could limit the access to other options, resulting in a balancing act. The subtheme Management represents the strategic activity of individuals to deal with limitations. Individuals look for achieving those options that have most value to them, despite the potential restrictions in access to other options (17, 18, 21-23).

> *“This morning, I got up—5 o’clock—I took my first pain killers, went back to bed again so that I was ready to get up to have my shower at half past six, or else, by the time you start taking them they haven’t taken effect and you’re trying to move around. So, yeah, you’ve got to think ahead…” (Female, not employed, A), Kinghorn, Robinson (17)*

Evaluation is a more fundamental theme. It represents the assessment of the realized options of an individual compared to his/her preferred options. This evaluation is influenced by the ability of individuals to access those preferred options. Moreover, the evaluation also is influenced by individuals’ expectations regarding the future ability to access those options (17, 19-21, 23). When the burden to access an option is extremely high, adaptation of preferences might occur. For example, a participant adapted her preferred options in the context of Activity Wellbeing, because she could not walk anymore.

> *“That would be a good point to put next to the hobbies, er walking…. I’m not able to do that, so then I had to look for something else to occupy my mind. So right, then—I always did do a lot of knitting and cross stitch—I take a lot of interest in that, and reading*.*” (Female, Retired), Kinghorn, Robinson (17)*

Some researchers suggest that an important prerequisite for adaptation is acceptance of the limitation to access (17, 23).

### Relationship between themes

The interaction between the limitations on the accessibility of preferred options, the ability of individuals to deal with these limitations, and the discrepancy between preferred level of accessing options and actual realized access of options, influenced how individuals perceived their level of control and their wellbeing. Limitations in access were experienced as a burden by the participants when they were unable to adapt their preferences (17, 22, 23). This burden particularly influenced the Emotional Wellbeing of individuals (18-20). Also, the results of the development papers suggest that participants experienced a loss of control when adaptation of preferences was difficult (17, 20, 23).

> *“…my health broke down again … which came as a shock… I had to give up work immediately …and it cast a long shadow because it’s always there in the background, you never know when it might jump on you. So you live with uncertainty*.*” (Female, 78), Al-Janabi, N Flynn (19)*

On the contrary, effectively managing and adapting to limitations could even produce a sense of pride (23). For instance, some participants considered themselves to be well off, even though limitations or burdens in access exist (17).

> *“I mean my discomfort is emotional rather than physical and I have days when I feel really good, and don’t worry, but if I have a very down period … I mean nobody wants to experience pain but I’m quite sure these days they can do things to relieve you of pain, but it’s just the emotional thing really which is more, especially when you’ve got nobody to talk it through with …” (Female, 83 years, PC) Sutton and Coast (21)*

### Post-Hoc sensitivity analysis

In respect to the COREQ checklist (14), four development papers did not provide information on all items (17, 19, 20, 22). The exclusion of these papers did not result in less or different themes emerging from the data, since all the themes and subthemes were identified in the three remaining papers. Nevertheless, excluding these four papers might have had an impact on the depth and transferability of the theoretical model.

## Discussion

Pettit’s theory (10) proved to be a useful a-priori “lens” to identify the differences between options, how these options are accessed, and how certain elements limit access to valued options. The application of the framework to the qualitative data meant that the a-priori themes were re-conceptualized. Furthermore, due to its clarity, the application of Pettit’s theory also facilitated the identification of elements that are important for wellbeing, but are not directly reflecting freedom itself.

The best-fit framework synthesis led to the re-conceptualization of the a-priori concept Access to Options to Perceived Access to Options. This change signifies that the participants reported their perceived ability to access options in the development papers. The a-priori concept Options was used to identify options that are important for individual. This re-conceptualized theme was called Option Wellbeing, with associated sub-themes that reflect various options that need to be fulfilled to a sufficient level for individuals to consider themselves to be well. One theme that emerged inductively from the data was Self-Realization, which reflects other elements related to living a meaningful life that are important for the experience of wellbeing. Finally, the theme Perceived Control emerged inductively from this analysis and reflects the capacity of the individuals to live with and adapt to blocks and burdens in access to options.

The theme Perceived Access to Options represents the perceived ability of individuals to realize options. Its subthemes reflect various sources of blocks and burdens that are linked to the level of fulfillment of the subthemes of Option Wellbeing. These reflect the burdens experienced by individuals that might hinder their ability to access options and consequently limit their capabilities. The assessment of those burdens is crucial to understand individuals’ wellbeing (see also Cookson (28) for a discussion in the context of the QALY). Consequently, as a theme it is more comprehensive than Sen’s conceptualization of capability in the way that it reflects (perceived) freedom. Thus, an instrument that aims to assess capabilities and is based on this theme should theoretically provide a large informational base.

Our analysis suggests another theme which might have a key role in wellbeing assessment: the theme Perceived Control. This theme represents the concept that peoples’ wellbeing is positively affected by the experience of being in control over their lives, even when there are restraints in access to options. The theme Perceived Control shows similarities with the concept of dis-capability of Bellanca, Biggeri (29). They suggest that an individual can be considered dis-capable when he or she is unable to manage the limitations that are imposed on her or his life due to, for example, a disability.

Two subthemes are linked to the theme Perceived Control. The first subtheme, Management, represents the idea that individuals use resources to deal with limitations in access. It also captures the fact that a constraint in access to a particular option could cause limitations in access to other capabilities, thus reflecting the strategic nature of control and coping. Previous empirical studies have already showed the importance of disease management for the wellbeing of the individuals. Gibbins, Bhatia (30) studied preferences of advanced cancer patients with chronic pain regarding pain management. Their result suggests that patients take just enough medication to have control over the limitations imposed by the pain, whilst minimizing the side effects of medication. They concluded that the extent to which pain and medication limits patients’ independence, understood as the range of choice and control patients have over their lives, is a stronger influence on the patients’ subjective wellbeing than the reduction in the experienced level of pain itself. Other studies show similar results concerning the importance of controlling the limitations that might be caused by disease (31), not only in respect to pain management (32, 33), but also in respect to controlling the symptoms of mental health problems (34).

The second subtheme of Perceived Control is Evaluation. It reflects the idea that individuals reevaluate the value of their options in relation to the limitations in access to those options. The aim of this reevaluation is to feel in control over their lives again and focus on new valuable options available to them. This mechanism is known as adaptation (31, 35, 36). In the context of patient wellbeing, facilitating adaptation to chronic disease can be seen as an integral part of treatment, particularly when no other possibilities to reduce symptoms exist (37).

Two further themes that emerged to be key for the assessment of wellbeing are the themes Self-Realization and Option Wellbeing. These two themes cover the experiences of individuals that are relevant for the assessment of wellbeing. The two themes are closely related to the way that subjective wellbeing is conceptualized (38). Subjective wellbeing has been conceptualized as consisting of two constructs: an “affect” construct, which reflects various types of emotions and a “cognitive” construct, which reflects experiences such as having a meaningful life. Furthermore, also the subthemes related to the themes Self-Realization and Option wellbeing show similarities to common abstract elements of wellbeing observed in different fields of study (39, 40). Examples of such commonalities are the importance of physical health or the importance self-determination (39, 40). Qizilbash (40) argues that one reason for these similarities across different fields of studies is that on a fundamental level these different lists of elements reflect commonly shared values.

Furthermore, he argues that differences between lists can be explained by the level on which they are applied (i.e. on the level of basic needs or on the level of capabilities). It is therefore not surprising that the themes and subthemes developed in this study reflect elements of wellbeing which have been cited to be important in various fields of study.

In the context of assessing the value of new health technologies, the present framework has two advantages over using Sen’s concept of capability as an informational base. First, it stresses that people who are able to access options *with* difficulty cannot be considered to have an equal level of capability compared to people who can achieve the same options *without* difficulty. Second, the framework expands the informational base of the capability approach to include the assessment of how people experience their capabilities in terms of the perceived control and its effect on the subjective wellbeing of individuals. Similar concerns have been raised by Clark (41), who argues that the informational base of the capability approach should be expanded to include a wider range of subjective experiences. Thus, the proposed framework provides a broad informational base on which to assess wellbeing. This in turn could lead to the refinement or development of instruments that assess the value of (medical) interventions.

### Limitations

The proposed framework is based on the synthesis of a limited number of qualitative papers. Furthermore, the synthesis depends on the interpretation of data by select number of researchers. Because of these limitations it is unclear how generalizable the framework is. Indeed, it is possible that some of the identified themes or subthemes are not as relevant as the researchers thought, whilst other potential themes might have been missed. Still, the themes seem to reflect common abstract elements which are important for wellbeing, given the parallels between the themes identified in this study and lists created by other authors.

## Data Availability

All data produced are available online at the cited articles.

## Notes

Conflict of interest: The authors declare that there are no conflicts of interest.

### Competing Interest Statement

The authors have declared no competing interest.

### Funding Statement

This study did not receive any funding

