## Appendix for "The value of freedom: extending the evaluative space of capability"

### Authors’ backgrounds

The data analysis itself was primarily conducted by the principal author, JU. JU has a background in health sciences and health economics. During the coding process, the analysis was discussed with EN, who provided a critical external view on the coding process and framework in development. EN is an anthropologist by training and has experience in conducting qualitative research in a variety of different settings. The results were discussed with KHV, who provided a critical view on the interpretation of the data and presentation of the results. KHV has a background in health economics and experience in economic evaluation. Because of the different backgrounds of the authors, the resulting discussion about the analysis created a richer and clearer framework.

### Access to options

Different takes exist on the nature of these blocks and burdens. When an option exists and can be accessed, then this combination counts as an option freedom. Pettit himself defines three distinct ways in which access to an option can be blocked for an individual: objectively, subjectively or both objectively and subjectively. An objective block to an option, for example, can be the lack of the bike lock key when an individual wants to access the option to ride a bike. In this case, the option freedom of an individual is blocked. A subjective block might occur when an individual has the possibility of cutting the bike lock with a bolt cutter. Some individuals might be afraid to be penalized when somebody notices that he/she is cutting the bike lock, while others are not afraid to get into trouble. In this case, the block is subjective, as there is nothing objectively stopping the individual to cut the bike lock.

Besides the possibility of access being blocked, Pettit also states the possibility of access being burdened. This means that an option is still accessible, but with difficulties. For example, when a bike is locked, an individual could bring the locked bike to a bike shop to cut the lock. This would cost considerable time and money, but the option to ride a bike can still be accessed. As such, the access to the option to ride a bike is not blocked, but burdened.

*Appendix table 1. Themes, subthemes and quotes from participants from development papers*

| themes | subthemes | description | Quotes |
| --- | --- | --- | --- |
| Option Wellbeing | Physical wellbeing | Physical wellbeing represents the physical health related wellbeing of an individual, such as the absence of discomfort or pain. | *“I just wouldn’t want to be in pain all the time.“ (Female, 72 years, PC), Sutton and Coast (23)*  *“I’m nearly 86, I’ve got all me faculties thank goodness, I saw my husband and he never ever knew me at times.” (Female, aged 85), Grewal, Lewis (22)* |
|  | Emotional wellbeing | Emotional wellbeing represents the affective wellbeing of an individual. Examples are feeling happy, or the absence of sadness or nervousness. | *“It’s contentment, I think, really—satisfaction. I love the theatre, I love the cinema ... if it’s just to go off for a day somewhere and have a meal in a pub… And I think that’s very essential, it’s just the simple pleasures of life really” (male, aged 69), Al-Janabi, N Flynn (21)*  *…obviously it’s [mother’s illness] been hard, it’s been upsetting…and visiting her now isn’t exactly a barrelful of laughs… I guess it’s saddening … (Female, 29), Al-Janabi, N Flynn (21)* |
|  | Social wellbeing | Social wellbeing represents the wellbeing associated with having good social contacts with, for example, friends or family. | *“One should take good care of the kids and the entire family, so that everyone is healthy and they can work properly and prosper." Greco, Skordis-Worrall (24)*  *“- you just feel so isolated” (Male, Employed), (19)* |
|  | Environmental wellbeing | Environmental wellbeing is related to the wellbeing associated with feeling settled in the house and the wider neighborhood. | *“A house should have a toilet, a bathing shelter, there should be a rubbish pit, and the house should be well taken care of. Even if you have all these things but they are not put to good use, diseases will be there.” Greco, Skordis-Worrall (24)*  *“I get a lot of pleasure out of sitting in the garden now... I suppose we are lucky living where we are… “(female, aged 73), (22)* |
|  | Activity wellbeing | Activity wellbeing represents the wellbeing derived from being able to do things for fun, or for relaxation. | *“Work is important. Just to go out and do things that aren’t mind numbing if you know what I mean.” (F employed), Kinghorn, Robinson (19)* |
| Reflective wellbeing | Having a role | Having a role represents wellbeing derived from the ability to do things that provide a sense of worth and is linked to the identity of individuals. | *“I do like playing…competitive sport…it’s got a bit of an edge …. I suppose through that there’s a bit of an achievement thing and it’s quite nice to be in a team or to be a captain for one of the teams” (Male, 29), Al-Janabi, N Flynn (21)*  *I’ve got four grandchildren, and I can’t pick none of them up, in fact, if they jump on me, I fall over. (Male, Not employed), Kinghorn, Robinson (19)*  *It brings a feeling of inner satisfaction really to think you have been of some use… Especially with bereavement, (male, aged 69), Grewal, Lewis (22)* |
|  | Having dignity | Having dignity is related to the wellbeing associated with being a respected member of a community, as well as being able to conduct as a being of worth. | *“A person who changes clothes is seen as living a good life. She changes dirty clothes after a bath, and puts on clean ones, and looks good. When she is amongst people, she is not shy. As for me, I may have to wash the few I have to put on when I go in public.” Greco, Skordis-Worrall (24)*  *I think some people get the wrong idea when they get these people who’ve got this Alzheimer’s that they wanna be directed and pushed – they don’t want that do they? (Female, 74 years, GP), Sutton and Coast (23)* |
|  | Being independent | Being independent represents the wellbeing derived from individuals being able to access options, without being interfered or having to rely on others. It is also linked to the perception of individuals to have a sense of agency, even though of problems that might rise up. | *“I would like to drive a bit more. Because I’m losing my independence. I have to rely on my husband to take me shopping now.” (Female, Retired), Kinghorn, Robinson (19)*  *“A person should be independent because when sick she doesn’t wait for someone to tell her what to do, men at times neglect that you are struggling.” Greco, Skordis-Worrall (24)*  *‘My family found it hard to believe their dad had Alzheimer’s... I said, ‘‘You heard what he called me a little while ago—‘Make me a cup of coffee, Nurse’’’. I said, ‘‘How do you think that makes me feel?’’ to my children.’ (female, aged 85), Grewal, Lewis (22)* |
|  | Self-determination | Self-determination represents the wellbeing that is a consequence of being able to make those choices that are meaningful to individuals. | *[Regarding the choice of conducting NIPT] “I just really think that women should be given ownership of the information and they can decide what they want to do.” Kibel and Vanstone (20)*  *“As a Physics Teacher, to do 6 years without any promotion is pretty unusual really because they’re in such short supply. And I was beginning to feel left on the shelf.” [Male, 28], Al-Janabi, N Flynn (21)* |
| Perceived access to options | Access due to physical wellbeing | Depending on the state of physical health, individuals have varying levels of access to options. | *[The chest infection] “just made it miserable for a week or two, I couldn’t get out or about…” [Male, 75], Al-Janabi, N Flynn (21)*  *“The negative thing with diabetes is when people at work ask if you can join them for something after work. No, I can’t, I’m going home to take my injection and have dinner. You get a little tied up, you know.” (#17; Woman, 60 years old, Type 2 DM), Engström, Leksell (25)*  *“Because of the arthritis and that, I can’t work, so there’s… work mates, you know, no Friday night when you’ve got the wages and you can enjoy it, a couple of beers. There’s none of that.” (Male, Not employed), Kinghorn, Robinson (19)* |
|  | Access due to emotional wellbeing | Also the emotional state of individuals can influence the access to options. People who do not feel well can experience difficulties accessing options. | *“I started getting depression… it’s like yesterday, I didn’t have a wash, I didn’t have a shave, I didn’t get up, I didn’t even unlock the door, and that was it” (Male, not employed, A), Kinghorn, Robinson (19)*  *“…if I feel fine up top I’ll have a better day. But if I wake up in the morning and I don’t feel right up here, you know the day is going to be worse“ (Male, not employed, B), Kinghorn, Robinson (19)* |
|  | Access due to social wellbeing | The social context of individuals also has an influence on the access of options of an individual. For example, having a wide support network eases the access the options. | *When I came out of hospital he [husband] done everything I mean, he cooked the food and he’s never cooked in his life [laugh] … And all the washing, ironing he did. (Female, 72 years, PC), Sutton and Coast (23)*  *“[Good life means] someone who when you meet on the way, can help you.”, Greco (30)*  *“I have many close friends and acquaintances who support me, which means a lot. Above all, in tough periods when it’s difficult to manage my blood glucose levels and so on, it’s a great support for me.” (#28; Man, 31 years old, Type 1 DM), Engström, Leksell (25)* |
|  | Access due to environmental wellbeing | The environment of an individual, inside and outside the house, also has an influence on the ease of access to options. An appropriate environment can facilitate the access to options. | *“I went in for this flat because it’s wheelchair friendly … I’m hoping that I’d lay here in a box, because it was a very deliberate act of me to look for somewhere where I can be independent for as long as possible.” (Female, 67 years, GP), Sutton and Coast (23)*  *It was lovely down here at one time, but it’s just frightening now… I wouldn’t walk when it were dusk from here to the top of the hill… because there’s weirdos on the canal. (female, aged 66), Grewal, Lewis (22)* |
|  | Access due to activity wellbeing | The activities of an individual influences the access to options. Due to an individual doing things, more options will come along the way. | *“Because of the arthritis and that, I can’t work, so there’s… work mates, you know, no Friday night when you’ve got the wages and you can enjoy it, a couple of beers. There’s none of that.” (Male, Not employed), Kinghorn, Robinson (19)*  *“At ante-natal classes …six of us really gelled and just became the closest of friends. It was like we’d known each other for years and years and years. … we see each other all of the time and we help each other out which is great.“ (Female, 32), Al-Janabi, N Flynn (21)* |
|  | Access due to financial resources | The financial situation of an individual has an influence on the ability to access options. The more financial resources an individuals has, the easier it is for an individual to access options. | *[Regarding the choice to do a NIPT] “If I had to pay for it, I would borrow from my friends or relatives. But I would just do anything possible to avoid a miscarriage.” Kibel and Vanstone (20)*  *“I’m reasonably fortunate… in so far as that we’ve got the two pensions… we’re able to go off... We grabbed a cheapie flight at the end of April... flew down to Nice...” (male, aged 70), Grewal, Lewis (22)* |
|  | Access due to technologies | The access to technologies gives the individuals the opportunity to access other options. For example, new drugs or therapies can take away limitations which are a consequence of disease. | *“I think the insulin pump is fantastic. Because it gives me freedom.” (#24; Woman, 64 years old, Type 1 DM), Engström, Leksell (25)*  *“if there was no NIPT, I'd be left with a positive result from my Integrated Prenatal Screening, not willing to do an amniocentesis because I'm not willing to you know, go with that risk, so the rest of my pregnancy I have that added stress of thinking that I have a high chance of having you know, a baby with Down Syndrome”, Kibel and Vanstone (20)* |
| Perceived control | Management | Management represents the perceived ability of individuals to manage the burdens or limitations in access to options. Being able to manage these burdens and limitations is conductive to the wellbeing of an individual. | *“This morning, I got up—5 o’clock—I took my first pain killers, went back to bed again so that I was ready to get up to have my shower at half past six, or else, by the time you start taking them they haven’t taken effect and you’re trying to move around. So, yeah, you’ve got to think ahead…” (Female, not employed, A), Kinghorn, Robinson (19)*  *[Regarding the management of diabetes] “It’s not easy, it’s an endless struggle to try to maintain good blood glucose levels. (...) It’s like walking a line.” (#24; Woman, 64 years old, Type 1 DM), Engström, Leksell (25)* |
|  | Evaluation | Evaluation represents the assessment by an individual between his or her realized options and the preferred level of option realization. | *“It is a constant sadness, that I’ve lost my sight... (...) But it’s nothing I get hung up on in my everyday life. (...) I consider myself as having a good quality of life.” (#1; Man, 49 years old, Type 1 DM), Engström, Leksell (25)*  *[Regarding NIPT] “I was going to go for the blood work but then I just was like, like you know what, I would rather not think about it, I would rather not stress about it.”, Kibel and Vanstone (20)* |

| No | Item | Page were item is addressed |
| --- | --- | --- |
| 1 | Aim | 6 |
| 2 | Synthesis methodology | 6 |
| 3 | Approach to searching | 7 |
| 4 | Inclusion criteria | 7 |
| 5 | Data sources | 7 |
| 6 | Electronic search strategy | Addressed in article by Ubels et al (9) |
| 7 | Study screening methods | Addressed in article by Ubels et al (9) |
| 8 | Study characteristics | Addressed in article by Ubels et al (9) |
| 9 | Study selection results | Addressed in article by Ubels et al (9) |
| 10 | Rationale for appraisal | Page 7 |
| 11 | Appraisal items | Page 7 |
| 12 | Appraisal process | Page 7 |
| 13 | Appraisal results | Page 8 |
| 14 | Data extraction | Page 8 |
| 15 | Software | Page 8 |
| 16 | Number of reviewers | Page 1 Appendix |
| 17 | Coding | Page 8 |
| 18 | Study comparison | Page 8 |
| 19 | Derivation of themes | Page 8 |
| 20 | Quotations | Page 8 |
| 21 | Synthesis output | Page 8 – 15 |

*Appendix table 2. ENTREQ checklist.*
